# Genome-Wide Association Study and Meta-Analysis of Phytosterols Identifies a Novel Locus for Serum Levels of Campesterol

**DOI:** 10.1101/2023.09.06.23295162

**Authors:** Jamil Alenbawi, Yasser A. Al-Sarraj, Umm-Kulthum I. Umlai, Ayat Kadhi, Nagham N. Hendi, Georges Nemer, Omar M. E. Albagha

## Abstract

**Background:** Sitosterolemia is a rare inherited disorder caused by mutations in the *ABCG5/ABCG8* genes. These genes encode proteins that are involved in the transport of plant sterols (phytosterols) out of the body. Mutations in these genes lead to decreased excretion of phytosterols, which can accumulate in the body and lead to a variety of health problems, including xanthoma formation, atherosclerosis, and premature coronary artery disease.

**Methods:** We conducted the first genome-wide association study (GWAS) in the Middle East/North Africa (MENA) population to identify genetic determinants of plant sterol levels in Qatari people. GWAS was performed on serum levels of β-sitosterol and campesterol measured using the Metabolon platform from the Qatar Biobank Cohort and genome sequence data provided by Qatar Genome Program. Furthermore, a trans-ancestry meta-analysis of data from our Qatari cohort with summary statistics from a previously published large cohort (9,758 subjects) of European ancestry was conducted.

**Results:** Using conditional analysis, we identified two independent single nucleotide polymorphisms (SNPs) associated with β-sitosterol (rs145164937 and rs4299376), and two independent SNPs associated with campesterol (rs7598542 and rs75901165) in the Qatari population in addition to two previously reported variants (rs199689137 and rs4245791). All of them map to the *ABCG5/8* locus except rs75901165 which is located within the Intraflagellar Transport 43 (*IFT43*) gene. The meta-analysis replicated most of the reported variants, and our study provided significant support for the association of variants in *SCARB1* and *ABO* with sitosterolemia.

**Conclusions:** This is the first study to shed light on genetic determinants of phytosterols in the MENA region using a genome-wide association approach. We also established the first polygenic risk score for this trait using the European large cohort study. These findings may have future implications on the personalized treatment of hyperlipidemia in general while showing the importance of building population-specific multi-omics databases.

**Clinical Perspective:**

- The formulation of the first polygenic risk score for sitosterolemia provides an exceptionally reliable tool for polygenic evaluation.
- Plant sterol measurement should be included in lipid panel checkups and genetic screening for patients with tendon xanthomas to ensure an accurate diagnosis.
- The establishment of regional and national registries for sitosterolemia in the Qatari community is essential for effective monitoring of the incidence of the disease and reducing the risk of early coronary artery disease.

## Introduction

Sitosterolemia (STSL1, OMIM #210250, STSL2, OMIM #618666) is an inherited metabolic disorder with an autosomal recessive pattern of inheritance. The condition is characterized by the accumulation of certain phytosterols, including sitosterol, campesterol, and stigmasterol, which are structurally similar to cholesterol and are commonly found in plant-derived foods. In individuals with sitosterolemia, these sterols are not effectively eliminated from the intestine or the liver, leading to their accumulation in various tissues and organs throughout the body. The aggregation of these sterols is believed to contribute to the development of cardiovascular disease and other related health complications. Sitosterolemia is a rare disorder, and its diagnosis requires specialized laboratory testing and genetic analysis ^(1, 2)^. The absorption process of dietary cholesterol and plant sterols is mediated by the Niemann-Pick C1-Like 1 (NPC1L1) transporter in the small intestine. It has been observed that the NPC1L1 transporter primarily absorbs dietary cholesterol, with plant sterols and stanols being absorbed to a lesser extent ^(3, 4)^. The efflux transporters, ABCG5 and ABCG8, have been identified as the main proteins responsible for the active transport of plant sterols back to the intestinal lumen ^(4, 5)^. The sterols are assimilated into chylomicrons that are rich in cholesterol and carried to the liver. These sterols then undergo another round of transportation by efflux through hepatic ABCG5/ABCG8 transporters ^(4, 6)^. Individuals affected by sitosterolemia may exhibit various clinical manifestations, including xanthomas, stomatocytosis, macrothrombocytopenia, splenomegaly, abnormal bleeding, and coronary artery disease (CAD). The presentation and severity of the condition can vary significantly, with some patients being completely asymptomatic while others develop severe symptoms and life-threatening complications. Therefore, early detection and management of sitosterolemia are critical to prevent or delay the onset of adverse outcomes associated with the condition (7–9). The STSL condition has been historically described as a rare disorder. However, recent research indicates that its frequency may be more prevalent than previously thought, with estimates suggesting a minimum occurrence of 1 in 50,000 individuals globally ^(10, 11)^. The differential diagnosis of STSL poses significant challenges due to the overlapping phenotypic features with other lipid disorders and the impracticability of conducting clinical testing of plasma phytosterols ^(12, 13)^. The confirmation of STSL diagnosis can be accomplished by detecting the levels of phytosterol in serum, which is widely regarded as the gold standard method. An even more optimal approach would be to utilize genetic testing to identify the most prevalent genes linked to STSL (14). One potential approach to accomplish this task would involve the identification of mutations in the genes encoding the ATP-binding cassette transporter (ABC transporters), specifically *ABCG5* and *ABCG8* ^(15, 16)^. The Genome Aggregation Database, accessible via the gnomAD browser v3.1.2 (https://gnomad.broadinstitute.org, accessed on 14^th^ of August 2023) has identified a total of 69 loss-of-function mutations in the *ABCG5* and *ABCG8*. These mutations have been shown to be associated with an increased risk of CAD (17–20). Furthermore, a mouse model of phytosterolemia exhibited cardiac damage and elevated mortality rates, despite achieving a 50% decrease in plasma cholesterol levels, according to a recent study (21). This finding highlights the complex and multifactorial nature of the cardiovascular disease and underscores the importance of exploring alternative mechanisms beyond traditional lipid-lowering therapies (21). The identification of the genetic susceptibility factors for STSL and its potential link to cardiovascular disease underscores the importance of further investigations. Currently, there have been only two genome-wide association studies (GWAS) conducted on STSL traits in European populations. The initial GWAS found that variants in *ABCG8* and the *ABO* locus were associated with increased levels of phytosterols and an increased risk of CAD (22). More recently, a genome-wide meta-analysis of phytosterols was performed to identify genetic loci associated with phytosterol levels and CAD in European populations (23). The authors conducted a genome-wide meta-analysis on 32 phytosterol characteristics to represent resorption, cholesterol synthesis, and esterification in six studies that included 9,758 participants. The study identified previously known associations between *ABCG5/8* and *ABO* and highlighted significant locus heterogeneity at *ABCG5/8* that influences several functional pathways. Additionally, novel loci, including *HMGCR*, *NPC1L1*, *PNLIPRP2*, *SCARB1*, and *APOE,* were discovered. The study employed Mendelian Randomization (MR) to establish a risk-increasing causal link between sitosterol blood concentrations and CAD, which was partially mediated by cholesterol. The authors concluded that phytosterols are polygenic and demonstrated that sitosterol has direct and indirect causal effects on CAD, which were further supported by MR analysis. Therefore, there is a critical need to expand upon the existing research and increase our understanding of the genetic underpinnings of STSL and its potential implications for cardiovascular health.

However, there is still a need for more evidence from genetic studies of sitosterolemia in Middle Eastern populations. Thus, we performed the first GWAS of estimated β-sitosterol and campesterol serum levels in the Middle Eastern populations from the Qatar Biobank (QBB) cohort. We identified a novel locus associated with campesterol on 14q24.3 (*IFT43*) and replicated Single Nucleotide Polymorphisms (SNPs) in the known loci at 2p21(*ABCG5/8*), 12q24.31(*SCARB1*) for β-sitosterol and 9q34.2, (*APO)* for campesterol. To further identify associated variants, we performed meta-analysis combining our findings with data from six population-based studies of European ancestry (N = 9,758 individuals) (23). Subsequently, we established a polygenic risk score to obtain further insights into the genetic architecture of this condition.

## Methods

### Study participants

The present study utilized data sourced from the QBB dataset, which is a unique population-based prospective cohort study consisting of Qatari nationals or long-term residents (who have been residing in Qatar for ≥ 15 years) who are over 18 years old. The QBB study collected physical measurements from all participants, and a standardized questionnaire was administered to obtain information regarding lifestyle and medical history. The study also conducted comprehensive baseline sociodemographic assessments, as well as clinical biomarkers and biochemical tests on the participants. The QBB project has been previously described in detail(24).

The study design for this research was approved by the Ethics Committee of the Hamad Medical Corporation (MOPH-AQBB-000222). Prior to participation, all participants in the QBB study signed an informed consent. This study was approved by the QBB Institutional Review Board (IRB) under Protocol no. E2021-QF-QBB-RES-ACC-00016-0158. To conduct the GWAS analysis, we utilized the first QBB dataset release, which included 6,218 individuals. To carry out the meta-analysis and replication analysis, we used a recently published European GWAS involving 9,758 individuals(23).

### QBB metabolomic plant sterols (phytosterols) profile measurement

The present study employed standard procedures for the collection of blood samples from all Qatari national participants, which were subsequently centrifuged to extract the serum fraction. The serum samples were then immediately stored at a temperature of -80°C. Untargeted metabolomics analysis was conducted on the collected serum samples utilizing established protocols. Metabolomic profiling was performed using the Metabolon HD4 technology, which employs ultrahigh-performance liquid chromatography and electrospray ionization tandem mass spectrometry to identify and quantify an expansive range of metabolites in each sample. The analytical approach enabled the detection and measurement of over a thousand metabolites in the sample, allowing for a comprehensive understanding of its metabolic composition.^(25, 26)^ Metabolomic measurements of plant sterols were obtained for β-sitosterol (n=1,652) and campesterol (n=1,451). Prior to conducting statistical analyses, a normalization step was performed on the phenotype through a rank-based inverse standard transformation using the R software -version 3.4.0 (www.r-project.org).

### Whole genome sequencing and subsequent bioinformatics analysis

The present study utilized whole genome sequencing and bioinformatics analysis to investigate genetic variations associated with serum levels of phytosterols. The genomic DNA was extracted from peripheral blood samples using an automated QIA Symphony SP instrument in accordance with the manufacturer’s instructions outlined in the Qiagen MIDI kit (Qiagen, Germany). The extracted DNA was quantified using the Quant-iT dsDNA assay (Invitrogen, USA) and the Flex Station 3 (Molecular Devices, USA). Whole genome sequencing was performed on the Illumina HiSeq X Ten (Illumina, USA) platform at the Sidra Clinical Genomics Laboratory Sequencing Facility. The sequencing was carried out with an average coverage of 30x, as previously described (27). This investigation employed Fast quality control (QC) data version 0.11.2 for the purpose of aligning sequences to the human reference genome GRCh38 via the Burrow-Wheeler Aligner version 7.12. Subsequently, variant calling was conducted using the Haplotype Caller, which was made available by the Genome Analysis Toolkit version 3.4. To generate a consolidated multi-sample Variant Call Format (VCF) file that encompasses all the samples, we performed joint calling on the individual genomic variant call files (gVCFs). This procedure comprised two stages: initially, Genomics DB was employed to unify regions across all the samples, and subsequently, Genotype VCFs (GVCFs) were utilized with SNP/indel recalibration to combine all the regions. Subsequently, in our downstream analysis, we exclusively considered variants that passed the GATK VQSR filtering criteria, with the PASS filter being our threshold for inclusion.

The present study conducted a comprehensive QC assessment using PLINK (version 2.0). SNPs with genotyping call rates less than 90%, minor allele frequencies (MAF) below 1%, or Hardy-Weinberg equilibrium *P-*values lower than 1.0×10^-6^ were excluded from the analysis. In addition, samples that did not meet the following criteria were excluded: call rate below 95% (N=1), duplicates (N=10), excess heterozygosity (N=8), and gender ambiguity (N=65). To further identify population ancestry outliers, we conducted a multidimensional scaling (mds) analysis using PLINK (27). We employed a pruned set of independent autosomal single nucleotide polymorphisms (SNPs), totaling 62,475, to construct a pairwise identity-by-state (IBS) matrix. To ensure that the population outliers were not included in the analysis, individuals who deviated from the mean of the first two multidimensional scaling (MDS) components by four or more standard deviation units (±4 SD) were excluded. Following the QC process, 6,047 subjects with clean WGS data were used but phenotype data was only available for 1,652 and 1,451 participants for β-sitosterol and campesterol, respectively, which were used in all downstream analyses. The QC process resulted in the availability of 8,218,470 variations, which served as the foundation for our GWAS.

### Genome-wide association analysis

We applied a generalized mixed model association test implemented in the Scalable and Accurate Implementation of Generalized mixed model (SAIGE) for genome-wide association testing, mainly in large biobank cohorts. We used SAIGE-v0.45 (28) to account for sample relatedness in single-variant association tests. Age, sex, total cholesterol levels, and the first four genetic principal components (PCs) were included as covariates in the regression model. R software -version 3.4.0 (www.r-project.org) was used to construct the genomic inflation factor, Quantile-Quantile graphs, and Manhattan plots (29). Furthermore, we used the GWAS catalog accessed on (on 11 June 2023) and PubMed literature searches to assess the relevance of the identified loci to Sitosterolemia and related phenotypes (30). Regional association maps were produced using the locusZoom tool and linkage disequilibrium data derived from the Qatari cohort (31). The threshold for genome-wide significance was *P* <5.0 ×10^-8^, and the threshold for suggestive evidence of association was *P* <1.0×10^-5^. For multiple association signals assessment, conditional association analyses were applied using SAIGE-v0.45. We then assessed loci reported in previous GWAS of Sitosterolemia for replication in our dataset. First, we evaluated the identified SNPs with robust genome-wide significant association with STSL from previous studies (*P* <5.0×10^-8^) (23). For SNPs that were not present in our dataset, we searched for close proxies (within ∼100 kb) in linkage disequilibrium (LD; r^2^>0.8) with the lead SNP based on LD data calculated from the Qatari population. We also evaluated SNPs associated with STSL (*P* <1.0×10^-5^) from the GWAS catalog for replication in our dataset using “Sitosterolemia, Sitosterol, phytosterolemia, and phytosterol” as phenotypes.

### Trans-ancestry meta-analysis

A trans-ancestry meta-analysis was conducted between the QGP data from the β-sitosterol and campesterol traits GWAS and summary statistics from the largest published GWAS of β-sitosterol and campesterol conducted by Scholz *et al.* (23) based on European ancestry participants. The β-sitosterol and campesterol meta-analyses allowed us to identify replicated SNPs as well as to identify novel variants or variants occurring in novel loci reaching the genome-wide significance level of *P* < 5.0×10^-8^. Summary statistics from the study of Scholz *et al.* were downloaded from the GWAS catalog and combined with our data in a fixed-effect and random-effect inverse variance meta-analysis using PLINK v1.9.The Cochran’s Q statistic was used to assess heterogeneity between studies in the meta-analysis.

### Validation of the findings from earlier studies with plant sterols

We compared the effect sizes and allele frequencies of the replicated variants from the Scholz *et al.* GWAS study. We further checked the GWAS catalog (December 12, 2022) for any previous reports of the genome-wide significant variants we identified via meta-analyses for β-sitosterol and campesterol to ascertain their novelty. Additionally, to check whether the β-sitosterol and campesterol QBB variants occurred in novel loci, we checked for genome-wide significant variants in the Scholz *et al*. GWAS study occurring within a +/-250kb region of our identified variants.

### Polygenic scores analysis

We accessed the Polygenic Score Catalog at https://www.PGSCatalog.org to investigate the availability of any Polygenic Risk Scores (PRS) related to sitosterol. However, we found that no such PRS was available. Consequently, we utilized whole-genome sequencing (WGS) data to assess the polygenic influence on sitosterol in our sample. To accomplish this, we calculated an SNP score (PRS) for each participant using a panel of six independent β-sitosterol SNPs identified by Scholz *et al.* (23). We employed linear regression models to examine the relationship between the levels of β-sitosterol in our dataset and polygenic risk score (PRS) while controlling variables of gender, age, total cholesterol levels, fasting time (hours) and PCs1-4. Specifically, the β-sitosterol single nucleotide polymorphism (SNP) scores for each participant were determined by computing the sum of the product of the effect size for the increasing allele of β-sitosterol (βx) and the number of increasing alleles (SNVx), which could be 0, 1, or 2 for each SNP (SNVx).

### SNP annotation

The present study investigated the relationship between identified plant sterol SNPs, specifically β-sitosterol and campesterol, and GWAS. To annotate these SNPs, we utilized Ensemble Variant Effect Predictor release 108 (VEP), which is available at https://grch38.ensembl.org/index.html(32). The present study utilized two publicly available databases, namely the Genome Aggregation Database (gnomAD) and the Allele Frequency Aggregator (ALFA), to perform a comparative analysis of the frequencies of the identified variants with those observed in the global population. The prevalence of the variants of interest across diverse populations was specifically investigated by accessing gnomAD https://gnomad.broadinstitute.org and ALFA www.ncbi.nlm.nih.gov/snp/docs/gsr/alfa/. Expression quantitative trait locus (eQTL) analysis was explored using the Genotype-Tissue Expression (GTEx) portal https://gtexportal.org/home/.

## Results

### Description of the study subjects

The cohort included 1,652 individuals with β-sitosterol levels available and 1,451 individuals with campesterol level measurements. Demographic analysis revealed that the cohort was evenly distributed in terms of gender, with 50.97% males and had a mean age of 39.7 years. Metabolomic data for plant sterols and laboratory measurements of lipid profiles, including total cholesterol, LDL cholesterol, HDL cholesterol, and triglycerides are presented in Table 1. We investigated the association between plant sterols and cholesterol levels in our study cohort. Correlation analysis between β-sitosterol and campesterol levels indicated a strong positive correlation between these two plant sterols (r(1645)=.87; P=2.2×10-6), as demonstrated in Figure 1-A. Furthermore, the correlation between serum levels of total cholesterol and campesterol (r(1209)= .33; P =4.2×10-38) or to β-sitosterol (r(1645)= .19; P =6.3×10-15) was found to be weak but significant, as shown in Figure 1-B and C.

**Figure 1:**
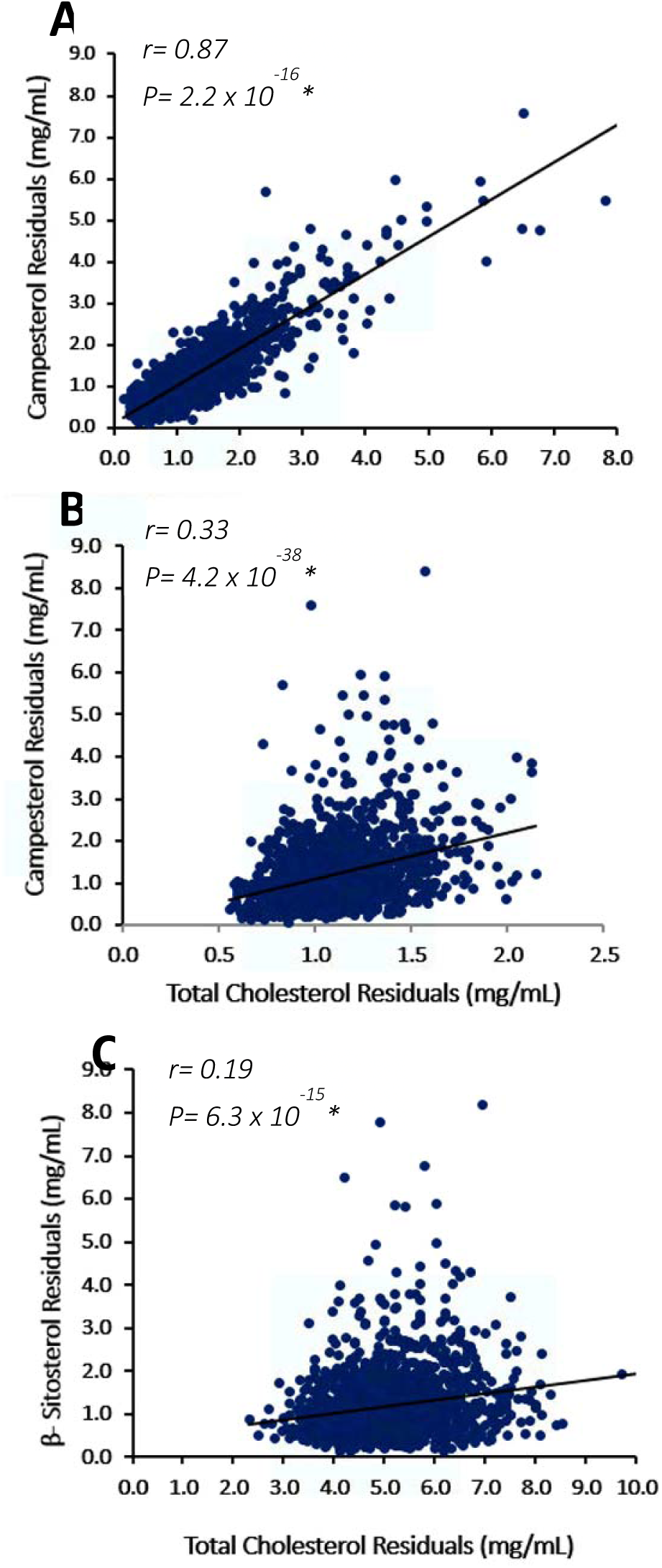
Correlation analysis between β-sitosterol, Campesterol, and Total Cholesterol. (A) Correlation plot for campesterol vs. 𝛽-sitosterol, (B) campesterol vs. total cholesterol, (C) β-sitosterol vs. total cholesterol. Pearson’s correlation analyses of β-sitosterol, campesterol, and Total Cholesterol were presented for quality control passed by participants of the QBB (n = 1,652; 1,451; 1,652), respectivelySignificant P-values are marked by asterisks and denote *P*-values less than 0.05. Abbreviations: R, Pearson’s correlation coefficient)

**Table 1.** Characteristics of the QBB study population.

|  | N | Mean<br>(mg/L) | M+4SD<br>(mg/L) | M-4SD<br>(mg/L) | Average age<br>(years) | Males | Females |
| --- | --- | --- | --- | --- | --- | --- | --- |
| **β- sitosterol** | 1,652 | 1.187 | 10.449 | -4.536 | 39.7 | 842 | 810 |
| **Campesterol** | 1,451 | 1.211 | 8.806 | -3.61 | 39.7 | 740 | 711 |

N: number of participants. Descriptive statistics are expressed as the average ( ± SD) or number (%) for QBB-quality participants. Values under columns “males” and “females” represents the number of participants.

### Genome-wide association study on β-sitosterol and campesterol traits

We utilized a genome-wide association approach and statistical fine-mapping analysis to investigate the genetic architecture of β-sitosterol and campesterol traits in the Qatari population of the Middle East. Our study included a total of 1,652 participants, with 1,451 individuals passing quality control. We focused our analysis on common and low-frequency risk alleles (MAF > 1%; N = 8,218,470) using linear mixed models to account for potential confounding factors, including age, sex, total cholesterol, and population principal-components. We employed quantile-quantile (Q-Q) and Manhattan plots to visualize the genetic associations for metabolomic β-sitosterol and campesterol trait concentrations (Figure 2). The Manhattan plots showed one strongly significant signal on 2p21 for both β-sitosterol and campesterol levels in the Qatari cohort, along another genome-wide significant signal on 14q24.3 for the campesterol trait only (Figure 2). Our results also indicated that the genomic inflation factor (λ_GC_) for β-sitosterol (λ_GC_ = 1.01) and campesterol (λ_GC_ = 0.99) showed no evidence of inflation (λ_GC_ = 1.01, 0.99), indicating no significant impact of population stratification or cryptic relatedness, as demonstrated by the Q-Q plot (Figure 2).

**Figure 2.**
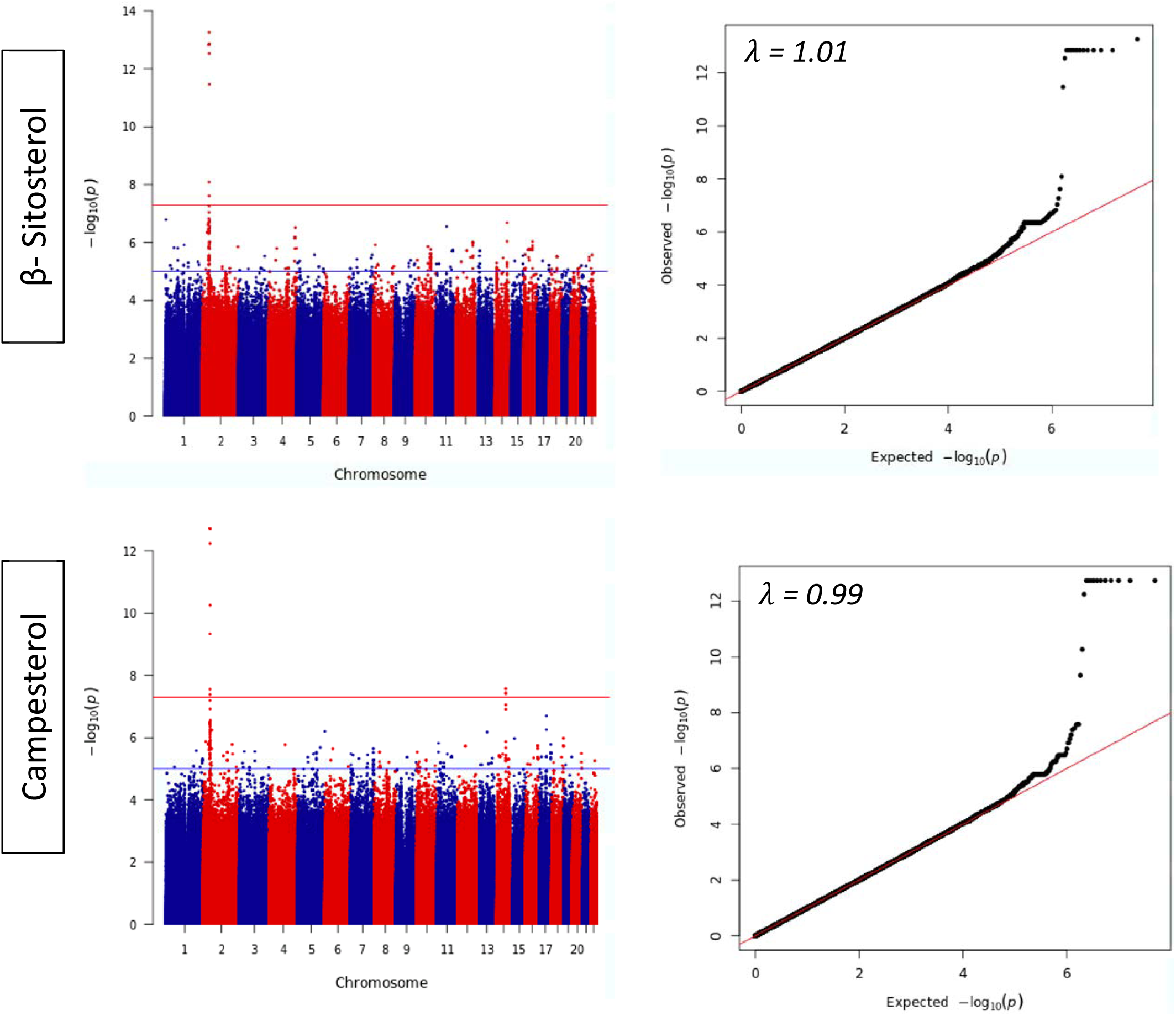
Genome-wide association results of metabolomic β-sitosterol & Campesterol levels were presented as Manhattan and Q-Q plots. The Manhattan plot illustrates the chromosomal positions of the genetic variants (N = 8,218,470) in relation to the –log10 *P*-value. The analysis was performed using linear mixed models that were adjusted for age, sex, total cholesterol, the first four principal components. The horizontal red line in the plot represents the genome-wide significance threshold (*P* ≤ 5.0 × 10^-8^), while a blue horizontal line indicates the threshold for suggestive evidence of association (*P*≤1.0 × 10^-5^). The Q-Q plot displays the observed –log10 *P*-values versus the expected –log10 *P*-values.

SNPs showing genome wide significant associations with β-sitosterol and campesterol are listed in Supplementary Table 1. Multiple genome-wide significant signals were observed on 2p21 for both β-sitosterol and campesterol; therefore, we conducted conditional analysis to identify independent signals from this locus (Supplementary Figures 1 and 2). For β-sitosterol, stepwise conditional analysis identified two independent genome-wide significant signals; rs145164937 *(P=*5.54×10^-14^) and rs4299376 *(P=*1.00×10^-12^) (Table 2) and two additional independent signals with suggestive evidence of association after conditional analysis (rs72796720; *P* =3.09×10^-6^, and rs781194036; *P=* 4.25×10^-5^) (Supplementary Table 2). The most significant markers were found within the genomic region containing *ABCG5/8*. Of these markers, only one was previously reported in the GWAS catalog as a genome-wide significant SNP, namely rs4299376, located in intron 2 of *ABCG8*. This SNP was significantly associated with the β-sitosterol trait in our cohort at a *P*-value of 1.0×10^-12^ (Table 2) and has been previously associated with the β-sitosterol trait in European populations(23). The other independent genome wide independent signal rs145164937 (Table 2) is a missense variant (p. Ala98Gly) that was not reported to be associated with β-sitosterol in the GWAS catalog. For the Campesterol trait, we identified two genome wide significant loci located on chr 2p21 and chr 14q24.3. The 2p21 locus has multiple genome wide significant signals spanning the *ABCG5/8* genes. Conditional analysis of this locus identified one independent signal (rs7598542) associated with campesterol levels (*P=* 4.15×10^-8^). As for the novel locus on chromosome 14q24.3 tagged by rs75901165 (*P=* 2.64×10^-8^) it maps within the *IFT43* gene (Table 2).

**Table 2.**
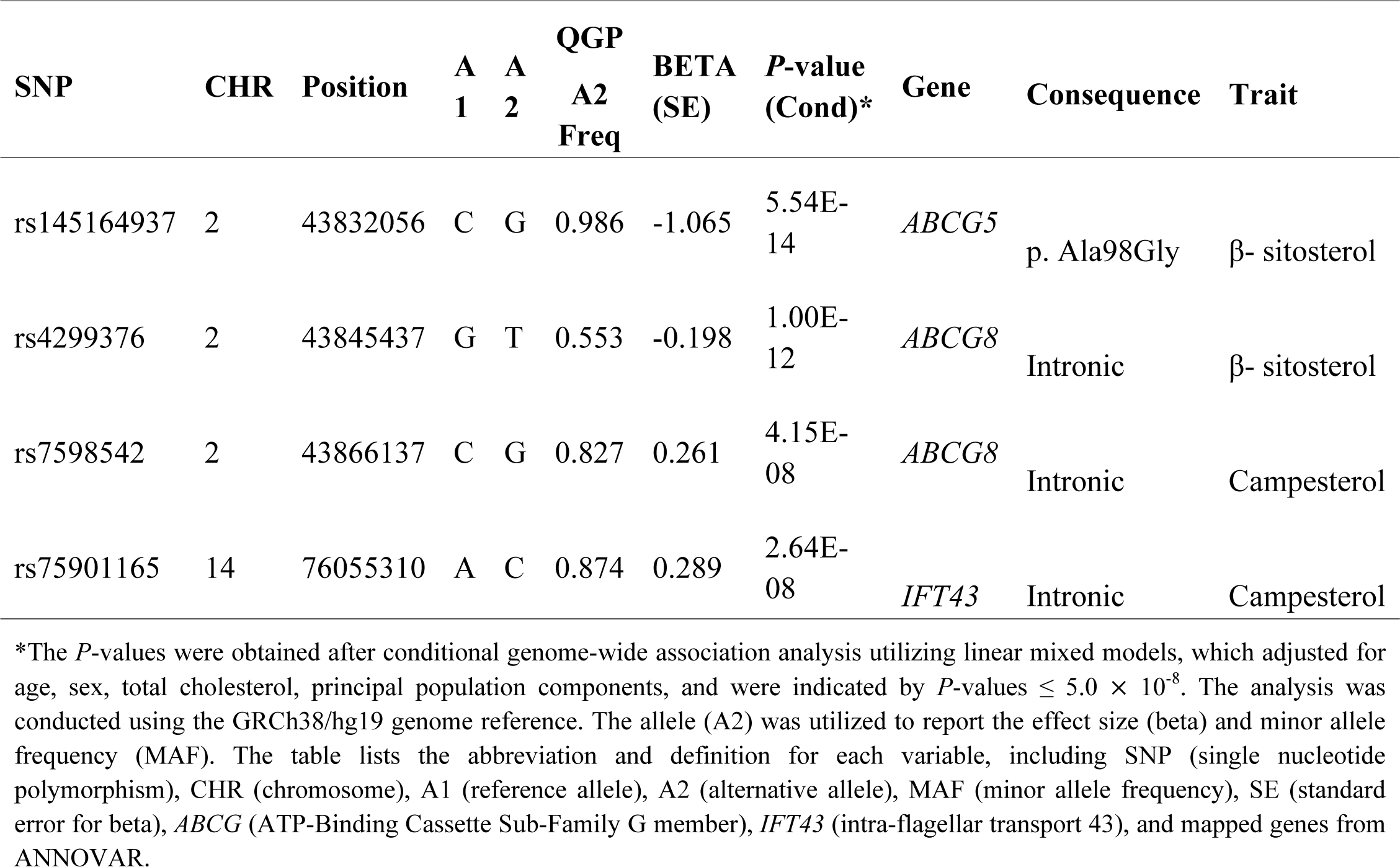
Independent Genome-Wide significant Variants Identified for β-Sitosterol & Campesterol.

| SNP | CHR | Position | A<br>1 | A<br>2 | QGP<br>A2<br>Freq | BETA<br>(SE) | P-value<br>(Cond)* | Gene | Consequence | Trait |
| --- | --- | --- | --- | --- | --- | --- | --- | --- | --- | --- |
| rs145164937 | 2 | 43832056 | C | G | 0.986 | -1.065 | 5.54E-14 | *ABCG5* | p. Ala98Gly | β- sitosterol |
| rs4299376 | 2 | 43845437 | G | T | 0.553 | -0.198 | 1.00E-12 | *ABCG8* | Intronic | β- sitosterol |
| rs7598542 | 2 | 43866137 | C | G | 0.827 | 0.261 | 4.15E-08 | *ABCG8* | Intronic | Campesterol |
| rs75901165 | 14 | 76055310 | A | C | 0.874 | 0.289 | 2.64E-08 | *IFT43* | Intronic | Campesterol |

\*The *P*-values were obtained after conditional genome-wide association analysis utilizing linear mixed models, which adjusted for age, sex, total cholesterol, principal population components, and were indicated by *P*-values $\leq 5.0 \times 10^{-8}$ . The analysis was conducted using the GRCh38/hg19 genome reference. The allele (A2) was utilized to report the effect size (beta) and minor allele frequency (MAF). The table lists the abbreviation and definition for each variable, including SNP (single nucleotide polymorphism), CHR (chromosome), A1 (reference allele), A2 (alternative allele), MAF (minor allele frequency), SE (standard error for beta), *ABCG* (ATP-Binding Cassette Sub-Family G member), *IFT43* (intra-flagellar transport 43), and mapped genes from ANNOVAR.

### Functional Expression

We further investigated the functional significance of the associated variants by evaluating expression quantitative trait loci (eQTL)) using the Genotype-Tissue Expression (GTEx) portal, Three SNPs (rs6544713, rs4245791, rs4299376) associated with β-sitosterol were found to be significantly associated with *ABCG8* gene expression in multiple tissues, but the highest significant association was in the large intestine (Figure 3). Interestingly, allele "C" in rs6544713 is strongly associated with increased ABCG8 expression (*P=*7.2×10^-16^), mainly in the transverse colon. Likewise, allele "T" in each of the rs4245791 (*P=*1.1×10^-16^) and rs4299376 *(P=*1.3×10^-15^) were significantly associated with ABCG8 elevated gene expression in the same tissue. (Figure 3-A, 3-B, & 3-C) respectively. The identified SNP (rs7598542) associated with campesterol trait showed that allele "C” is also significantly associated with increased ABCG8 expression with highest significance (*P=*4.3×10^-7^) in the colon amongst all other tissues (Figure 3-D).

**Figure 3:**
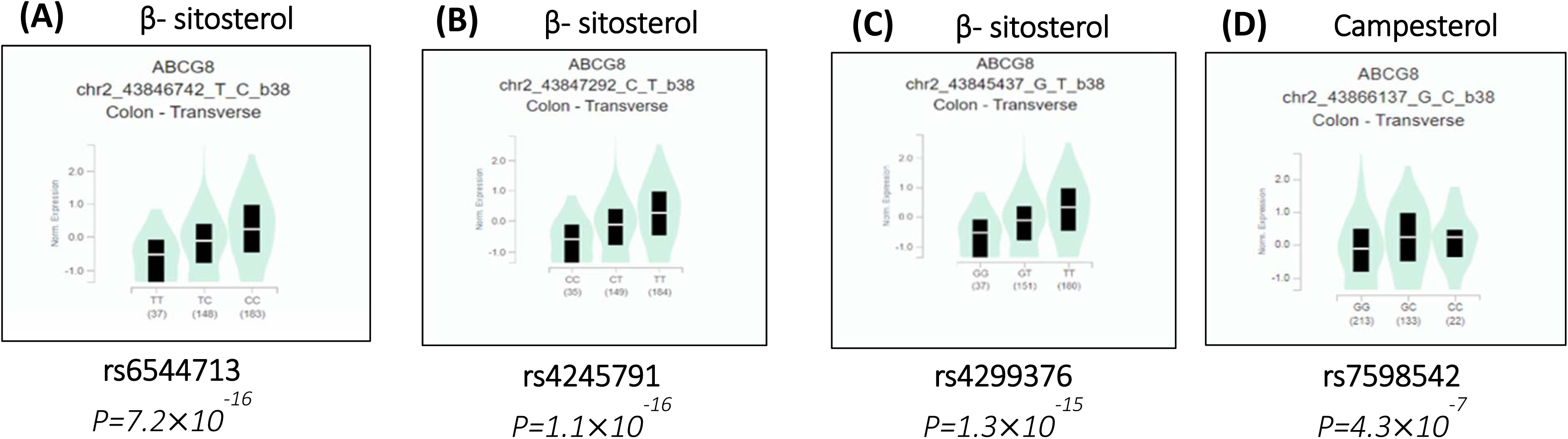
Single-Tissue Violin Plots. Gene expression of the associated variants (expression quantitative trait loci (eQTL)) using the Genotype-Tissue Expression (GTEx) portal. The data Source: GTEx Analysis Release V8 (dbGaP Accession phs000424.v8. p2). (A) rs6544713 (chr2_43846742_T_C). (B) rs4245791 (chr2_43847292_C_T). (C) rs4299376 (chr2_43845437_G_T). (D) rs7598542 (chr2_43866137_G_C)

**Figure 4.**
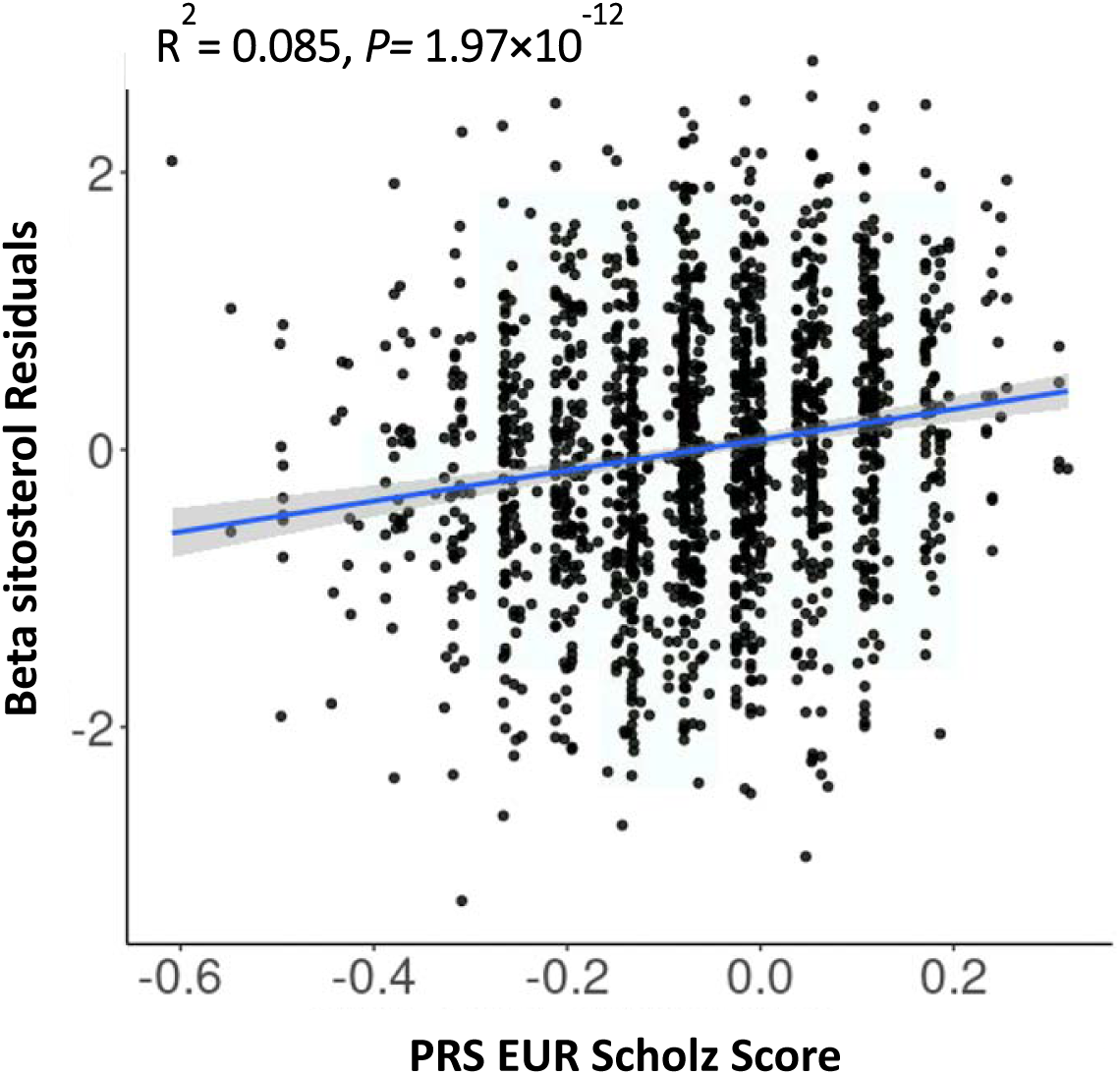
The performance of polygenic scores calculated from the European study on the Qatari population. To evaluate the effectiveness of polygenic scores obtained from a European study on the Qatari population. Specifically, we examined the relationship between inverse-normalized baseline levels of β-sitosterol and weighted polygenic risk scores (PRS) derived from the Scholz *et al.* study. Regression analysis was conducted to assess this correlation, with the blue line representing the most suitable fit.

### Replication of Known Loci

We assessed the extent of replication of previously published work on Sitosterolemia. Our study nominally replicated (*P-*value ≤ 0.05) most of the loci identified in Scholz *et al.* (Supplementary Table 3). One of the replicated loci, rs4299376, which reached the GWAS threshold in Scholz *et al.,* was also significantly replicated *(P*-value < 5.0×10^-8^) in our study. Next, we investigated the correlation between effect directions and effect size for the replicated SNPs at the genome wide significance threshold (*P*-value < 5.0×10^-8^) or nominally replicated (*P-*value ≤ 0.05) for β-sitosterol and campesterol traits. All the replicated SNPs showed consistent directionality, with some variation in the effect sizes compared to those in the study by Scholz *et al.* (Supplementary Table 3).

### Trans-ancestry Meta-analysis for Sitosterolemia

In order to increase the statistical power for identifying potential novel variants, we employed a meta-analysis approach. Specifically, we combined our GWAS data with the European GWAS study by Scholz *et al.* (23). Our meta-analysis validated the replication of several variants for β-sitosterol (n=166) and campesterol (n= 142) traits (Supplementary Table 4 & 5 respectively). Of the 166 replicated variants in the β-sitosterol trait, five variants; rs10846742 (*P* = 9.39×10^-9^), rs7485656 (*P*=1.37×10^-9^), rs4148221(*P*=5.22×10^-9^), rs150144845*(P*=1.34×10^-10^), rs112403212 (*P*= 1.26 ×10^-9^), were initially below the genome significance threshold in the Scholz *et al.* study but reached the significance threshold after combining with the QGP cohort. Among these five variants, two are located withing the *ABCG5/8* locus (rs150144845 in *ABCG5* and rs4148221 in *ABCG8*) whereas the remaining within *SCARB1* (rs10846742, rs7485656, and rs112403212). This locus was only tagged by one SNP (rs10846744) in the Scholz *et al.* study, and meta-analysis results provided additional evidence for the association of *SCARB1 with* β-sitosterol, which is a critical component in the reverse cholesterol transport pathway and plays a crucial role in lipid metabolism (Supplementary Table 6). Furthermore, out of the 142 replicated variants in the campesterol trait, two variants rs600038 (*P* = 8.2 ×10^-9^) and rs651007 (*P* = 8.55×10^-9^) *ABO*, were of borderline genome significance in the Scholz *et al.* study but reached the genome-wide significance threshold after combining with the QGP cohort. These loci were only tagged by one SNP (rs2519093) in *ABO* in the Scholz *et al.* study. The findings from the meta-analysis of both the discovery and replication cohorts are presented in Supplementary Table 7.

### Estimation of the sitosterolemia polygenic score

We used a panel consisting of six independent β-Sitosterol SNPs, as identified by Scholz *et al.* (23) to determine the polygenic score (PRS) of the QGP participants and evaluate its performance at predicting serum levels of β-Sitosterol. We then examined a linear regression model for the fasting and non-fasting data; using predictors: PRS score, gender, age, total cholesterol levels, fasting time (hours) and PCs1-4. Goodness of model fit indicated an R^2^= 0.089 (adjusted-R^2^= 0.083) for fasting and R2= 0.085 ((adjusted-R^2^= 0.082) for non-fasting with significant P-values for the PRS scores in the model with P=1.28×10^-12^ and 1.97×10^-12^ respectively.

## Discussion

The study analysed the contribution of genetic variation to metabolomic measurements of plant sterols within a subset of 1,652 individuals for β-sitosterol and 1,451 individuals for campesterol measurements from a larger cohort of 6,218 Qatari participants. It is only the second such study worldwide, aimed at identifying genomic loci associated with sitosterolemia, a rare inherited disease, which can be misdiagnosed and/or masked by familial hypercholesterolemia.

### *ABCG5/8* are the major players across populations for both beta-sitosterol and campesterol

Our results showed that 18 and 20 variants were significantly associated with β-sitosterol and campesterol traits, respectively, at the genome-wide significance level of *P*<5.0×10^-8^. Notably, our findings confirmed that the *ABCG5/8* and 2p21 locus were the primary predictors of sitosterol, with three out of four independent variants mapped within this region. At the 2p21 locus, we have confirmed a significant association with sitosterol, consistent with previous research (33). Specifically, the results revealed that four independent SNPs, namely rs145164937 and rs4299376 for the β-sitosterol trait, and rs7598542, and rs75901165 for the campesterol trait, reached genome-wide significant associations (*P*-value < 5.0×10^-8^), while two independent SNPs, namely rs72796720, rs781194036 had suggestive associations for β-sitosterol trait (*P*-value < 1.0×10^-5^). All are novel SNPs except for rs4299376, which had been previously reported in the GWAS Catalog by Scholz *et al.* where the unconditional analysis associated it to the trait with a *P-value* of 9.5x10^-74^ (23). Despite being mapped to intron 2 of *ABCG8* with no clinical variant attributed in the ClinVar database, this SNP is one of the six independent variants derived from the European study that we used to devise our polygenic risk score. The allele frequency of allele 2 (T) of the variant in the QGP cohort is 0.553, whereas it was 0.68 in the Scholz study and 0.757 in the gnomAD. This highlights the slight differences between populations albeit the significant differences between the cohorts’ size. Functionally, the phenotypic measurements of plant sterol revealed that there are 774 individuals with the heterozygous variant, exhibiting an average β-sitosterol of 1.18, and 523 individuals with the homozygous variant, exhibiting an average β-sitosterol of 1.06. This was corroborated by eQTL analysis of the two alleles in human tissues, specifically in the colon whereby the TT genotype is correlated with higher expression of the transporter (Figure 3).

The unique features related to the Qatari population were mostly highlighted by the identification of the other 3 novel SNPs in the same locus, in particular the rs145164937 variant. ClinVar shows conflicting interpretation of pathogenicity for this SNP. It is a novel finding as it has not been reported in the GWAS Catalog. Functionally, rs145164937 leads to an amino acid change in the N-terminal part of the protein (p.Gly98Ala) near the functional region implicated in binding ATP (Walker A). The QGP allele frequency for allele “C” (encoding Alanine) is 0.01, whereas its frequency in Europeans is 0.002 (gnomAD. A significant *P*-value of 5.54×10^-14^ is observed, indicating the association between the mutation and the β-sitosterol levels. Furthermore, the effect direction for the C allele (Beta=+1.065), indicating that the presence of Alanine could leads to a decrease in ABCG5 transporter function, and potentially leading to increased level of serum β-sitosterol. This reduced functionality was also reported in a large European cohort study including the UK biobank. The results showed that rs145164937 “C” allele was associated with increased risk of CAD (18).

The measurement of all plant sterols (specially the β-sitosterol) using metabolomics is crucial for diagnosing Sitosterolemia, which represents an important diagnostic criterion and is the predominant form of sterol found in Sitosterolemia patients (34). In this study, we aimed to identify genetic variants associated with β-sitosterol and campesterol traits using genome-wide association analysis. Overall, our results provide new insights into the genetic architecture of sitosterol metabolism and may have implications for the diagnosis and treatment of sitosterolemia.

### Expanding the genetic spectrum: *SCARB1* and *IFT43* have distinctive signatures

While findings of the major role played by the ABCG5/8 were concordant for both campesterol and beta-sitosterol, small and significant divergence was found in two loci encoding the *SCARB1* and *IFT43* genes. Our study did consolidate the previous European findings on the novel role of *SCARB1* by listing 3 variants (rs10846742, rs7485656, and rs112403212) as opposed to only one (rs10846744) by the previous study. While the functional role of the SCARB1 protein in sitosterol metabolism is not known, its role in reverse cholesterol transport is well documented highlighting the intimate relationship between plant and animal sterols in terms of absorption and excretion from the human body. The findings suggest that the addition of the new data provided stronger evidence for the association of *SCARB1* with the trait. *SCARB1* is a crucial component in the reverse cholesterol transport pathway and plays an essential role in lipid metabolism.

The rs75901165 variant on chr14-(76055310) maps to an intronic region in the Intraflagellar transport (*IFT*) gene. The *IFT43* gene has been implicated in the regulation of cilium length and IFT (35). Mutations in *IFT43* have been associated with ciliopathies, including skeletal (36) However, none of the previous studies involving human subjects had assessed associations with sterols. Our investigation yielded no association of this variant or any other variant within the *IFT43* in the European cohort. Interestingly, a knock-out of this gene is embryonically lethal, but the heterozygous mice showed a statistically significant increase in circulating cholesterol level with a *P*-value of 1×10^-04^.(<u>Ift43 Mouse Gene Details | intraflagellar transport 43 |</u> <u>International Mouse Phenotyping Consortium</u> (mousephenotype.org). Additional functional assays are needed to ascertain a role for *IFT43* in sitosterolemia while a more extensive cohort is needed from the MENA region to confirm the genomics findings and cross-compared to other populations.

Whether the differences obtained between the two traits relate to the function of each sterol in terms of its chemical structure and mode of absorption and elimination is to be further studied using in vitro and in vivo mice models. Of note that in humans the hepatic clearance of campesterol is significant lower than that of beta-sitosterol (37), whereas it is stigmasterol accumulation and not the other two plant sterols that causes cardiac injury in mice (21)

### Developing a PGS for early diagnosis

This study aimed to develop a PRS for sitosterolaemia. Due to the limited sample size, we were unable to construct the PRS based on identified independent variants and test it on the same data. Instead, we utilized the six independent sitosterol SNPs identified by Scholz *et al.* to calculate the PRS. Our findings revealed a positive model fit of R^2^= 0.085 (adjusted-R^2^=0.081, P=1.97x10^-12^, df= 1638) and significant correlation (r= 0.17, df = 1645, P = 6.3×10^-12^) between β-sitosterol levels in the samples and the calculated PRS. While the significance of such a score is excellent, its application on other populations needs to be tested and other variants in specific populations like the IFT43 one in the Qatari population could add more accuracy to the results.

In conclusion, our study has identified four independent variants in the GWAS that are associated with sitosterolemia in the Qatari population. Our results provide new insights into the genetic architecture of sitosterol metabolism and may have implications for the diagnosis and treatment of sitosterolemia. Our findings serve as a foundational study for future comparative research in this area.

## Supplementary Information

Supplementary Table 1

Supplementary Table 2

Supplementary Table 3

Supplementary Table 4

Supplementary Table 5

Supplementary Table 6

Supplementary Table 7

Supplementary Figure 1

Supplementary Figure 2

## Availability of code

The analysis was carried out utilizing openly accessible software instruments that have been referenced in both the primary text and the Methodology segment.

## Author contribution

The study was conceptualized, and the model was designed by JA, OA, and GN. YA was responsible for creating the codes used in the analysis of GWAS and meta-analyses, while YA and UU were responsible for calculating the polygenic risk score and conducting the conditional analysis. JA carried out the data analysis and interpretation of the results and also prepared the initial draft. AK, N.N.H and all the authors contributed to the final manuscript writing and reached a consensus on its content.

## Ethics declarations

### Competing interests

It is hereby declared that the authors have no conflicts of interest to disclose.

## Data Availability

The data used in this study are subject to certain licenses and restrictions. The raw whole-genome sequence data from Qatar Biobank cannot be deposited into public databases due to data privacy laws. However, access to the QBB/QGP phenotype and whole-genome sequence data can be obtained through an ISO-certified protocol. This involves submitting a project request at https://www.qatarbiobank.org.qa/research/how-apply, which must be approved by the Institutional Review Board of the QBB. If you wish to access these datasets, please visit https://www.qatarbiobank.org.qa/research/how-apply for more information.

## Acknowledgements

J.M.A was supported by a Ph.D. scholarship from Hamad Bin Khalifa University (HBKU) funded by the Qatar Foundation. The Qatar Biobank (QBB) and Qatar Genome Program (QGP) are Research, Development, and Innovation entities within the Qatar Foundation for Education, Science, and Community Development.

## Funding

Open access funding provided by the Qatar National Library.

## Author information

Authors and Affiliations

## Notes

### Competing Interest Statement

The authors have declared no competing interest.

### Clinical Trial

No clinical trial.

### Funding Statement

No external funding was recieved.

### Author Declarations

This study was approved by the QBB Institutional Review Board (IRB) under Protocol no. E2021-QF-QBB-RES-ACC-00016-0158.

